# Cancer is associated with the severity and mortality of patients with COVID-19: a systematic review and meta-analysis

**DOI:** 10.1101/2020.05.01.20087031

**Authors:** Ya Gao, Ming Liu, Shuzhen Shi, Yamin Chen, Yue Sun, Ji Chen, Jinhui Tian

**Affiliations:** Evidence-Based Medicine Center, School of Basic Medical Sciences, Lanzhou University, Lanzhou 730000, China; Evidence-Based Nursing Center, School of Nursing, Lanzhou University, Lanzhou 730000, China; Key Laboratory of Evidence-based Medicine and Knowledge Translation of Gansu Province, Lanzhou University, Lanzhou 730000, China

**Keywords:** COVID-19, Neoplasms, Prevalence, Severe illness, Prognosis, Meta-analysis

## Abstract

**Background:** Cancer patients are considered a highly vulnerable population in the COVID-19 epidemic, but the relationship between cancer and the severity and mortality of patients with COVID-19 remains unclear. This study aimed to explore the prevalence of cancer in patients with COVID-19 and to examine whether cancer patients with COVID-19 may be at an increased risk of severe illness and mortality.

**Methods:** A comprehensive electronic search in seven databases was performed, to identified studies reporting the prevalence of cancer in COVID-19 patients, or providing data of cancer between patients with severe or non-severe illness or between non-survivors and survivors. Meta-analyses were performed to estimate the pooled prevalence and odds risk (OR) using the inverse variance method with the random-effects model.

**Results:** Thirty-four studies with 8080 patients were included. The pooled prevalence of cancer in patients with COVID-19 was 2.0% (95% CI: 2.0% to 3.0%). The prevalence in Italy (5.0%), France (6.0%), and Korea (4.0%) were higher than that in China (2.0%). Cancer was associated with a 2.84-fold significantly increased risk of severe illness (OR = 2.84, 95%CI: 1.75 to 4.62, *P* < 0.001) and a 2.60-fold increased risk of death (OR = 2.60, 95%CI: 1.28 to 5.26, *P* = 0.008) in patients with COVID-19. Sensitivity analyses showed that the results were stable after excluding studies with a sample size of less than 100.

**Conclusions:** Cancer patients have an increased risk of COVID-19 and cancer was associated with a significantly increased risk of severity and mortality of patients with COVID-19.

## 1. Introduction

Coronavirus disease 2019 (COVID-19), acute pneumonia caused by severe acute respiratory syndrome coronavirus 2 (SARS-CoV-2) infection, first appeared in December 2019 and rapidly spread to a large number of countries and has become a pandemic [1–3]. As of April 22, 2020, a total of 2,471,136 laboratory-confirmed cases were reported worldwide, with a mortality rate of 6.8% [4]. Previous studies have shown that comorbidities in patients with COVID-19 are associated with poor prognosis, including hypertension, diabetes, chronic obstructive pulmonary disease (COPD), cardiovascular disease, and cerebrovascular disease [5–7].

Cancer patients are in a state of systemic immunosuppression and are considered a highly vulnerable population in the COVID-19 epidemic [8, 9]. Previous original studies revealed that cancer patients infected with COVID-19 had a higher risk of serious clinical events and death than those without cancer [9, 10]. A previous meta-analysis also evaluated the relationship between cancer and patients with COVID-19 and suggested cancer did not increase the risk of disease progression. Another two meta-analyses investigated the prevalence of cancer among patients with COVID-19, but their results were inconsistent. Furthermore, these meta-analyses were limited by the small sample size and the conclusions were inconclusive. Therefore, a comprehensive meta-analysis is urgently needed to answer clinical questions. The primary objective of this study was to investigate the prevalence of cancer in patients with COVID-19. The secondary objectives were to examine whether cancer patients with COVID-19 may be at an increased risk of severe illness and whether cancer is associated with mortality in patients with COVID-19.

## 2. Methods

We reported this systematic review following the Preferred Reporting Items for Systematic Reviews and Meta-Analyses (PRISMA) statement [11]. The study protocol has been registered in the International Prospective Register of Systematic Reviews (PROSPERO, CRD42020181622).

### 2.1. Eligibility criteria

Studies that met the following criteria were included: (1) patients have a laboratory-confirmed diagnosis of COVID-19; (2) described the prevalence of cancer in infected patients, or provided data of cancer between patients with severe or non-severe illness or between non-survivors and survivors; (3) published in Chinese and English.

We excluded following studies: (1) studies with a sample size of fewer than 20 patients; (2) studies focused on only suspected cases or suspected cases and confirmed cases; (3) studies did report data related to cancer patients; (4) review articles, protocols, guidelines, consensus, comments, abstracts, letters, and editorials.

### 2.2. Literature search

We conducted comprehensive searches in PubMed, EMBASE.com, the Cochrane Central Register of Controlled Trials (CENTRAL), Web of Science, Chinese Biomedical Literature Database (CBM), China National Knowledge Infrastructure (CNKI), and Wanfang Database up to April 12, 2020. Search terms included the following words: “COVID-19”, “coronavirus disease-19”, “new coronavirus”, “2019-nCoV”, “novel corona virus”, “novel coronavirus”, “nCoV-2019”, “novel coronavirus pneumonia”, “2019 novel coronavirus”, “coronavirus disease 2019”, “SARS-CoV-2”, “severe acute respiratory syndrome coronavirus 2”, “neoplasms”, “neoplasia”, “tumor”, “tumour”, “cancer”, “malignancy”, “clinical characteristic” “clinical feature”, “risk factor”, and “comorbidities”. The search strategy of PubMed is presented in Appendix Word 1. Reference lists of eligible studies and relevant systematic reviews were manually searched for potentially eligible studies.

### 2.3. Study selection process

The retrieved records were imported into EndNote X8 (Thomson Reuters (Scientific) LLC Philadelphia, PA, US) software for management. Two authors independently (YG and ML) screened the titles and abstracts of the records to determine if they met the inclusion criteria. Then, the same two authors retrieved the full text of all potentially eligible studies and assessed the eligibility of each study according to the inclusion criteria. Regarding multiple studies from the same teams or studies with samples from the same settings, we evaluated the time frame and detailed data of the study. For studies with overlapping data, we included the study with a larger sample size. Conflicts were resolved by discussions with a third reviewer (JHT).

### 2.4. Data extraction and quality assessment

A standardized data extraction form was developed using Microsoft Excel 2016 (Microsoft Corp, Redmond, WA, www.microsoft.com) through discussions with the review team and was revised after piloting on a random of five studies. The data extracted included: (1) study characteristics (first author, year of publication, country of the corresponding author, journal name, publication language, study setting, recruitment time frame), population characteristics (age, sex, sample size), and outcomes of interest (number of cancer patients, sever cases, non-severe cases, deaths, and survivors). The severe illness was defined in this study as patients experiencing acute respiratory distress syndrome (ARDS), requiring mechanical ventilation, requiring vital life support, or requiring intensive care unit admission (ICU) support [12, 13]. We used the Newcastle-Ottawa quality assessment scale (NOS) to assess the quality of included studies [14]. Studies that obtained more than 7 stars were considered as high quality, 5-7 stars were considered as moderate quality, and lower than 5 stars were considered as low quality [15]. The data extraction and quality assessment were performed by one reviewer (YG, ML, SZS, or YMC) and checked by a second reviewer (YS and JC). Discrepancies were resolved by consensus or by the discussion with a third reviewer (JHT).

### 2.5. Statistical analysis

We conducted a rate meta-analysis to estimate the pooled prevalence and its 95% confidence interval (CI) for cancer among patients with COVID-19. Pairwise meta-analyses were conducted to compute the odds ratio (OR) and 95% CI of cancer prevalence in COVID-19 patients with or without severe illness, and non-survivors or survivors. The meta-analyses were performed using the inverse variance method with the random-effects model. The statistical heterogeneity was assessed with the I^2^ statistic, and value of < 25%, 26-50%, and > 50% was considered as low, moderate, and high level of heterogeneity, respectively [16].

We planned to conduct subgroup analyses of the outcomes between different countries. Sensitivity analyses were conducted by excluding studies with a sample size of less than 100. We also performed univariate meta-regression analyses to assess if either the outcomes or the heterogeneity was associated with the publication languages and number of centers of the study conducted. The funnel plot and Eggers test were adopted to detect the publication bias. All analyses were conducted using Stata (13.0; Stata Corporation, College Station, Texas, USA Stata) and the statistical level of significance was set at *P* < 0.05.

## 3. Results

### 3.1. Screening results

We identified 1592 records through the literature search, among which 838 were from English databases, 751 were from Chinese databases, and 3 were from other sources. After removing duplicates and reviewing the titles and abstracts, 1466 records were excluded. Through full-text evaluation of the remaining 126 records, 92 records were further excluded. Finally, 34 studies [17–50] were included for our meta-analyses. The flowchart of the screening process is presented in Figure 1.

**Figure 1.**
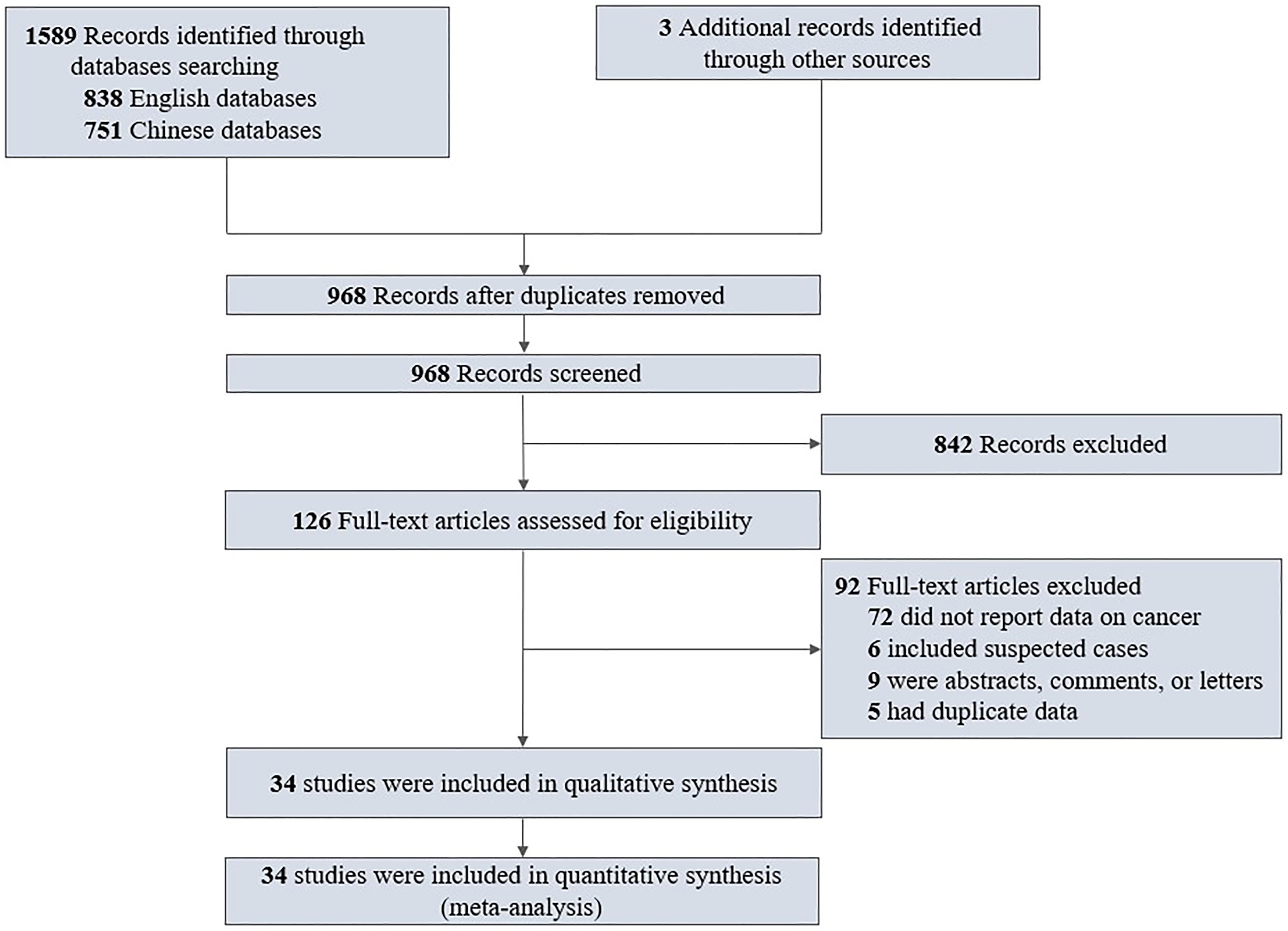
The flowchart of the screening process

### 3.2. General characteristics and quality of studies

All included studies [17–50] were published in 2020, enrolled patients between December 11, 2019 and March 21, 2020. 31 studies [17–20, 23–25, 27–50] were performed in China and 3 studies [21, 22, 26] were conducted in Italy, France, and Korea. 23 studies [17–39] published in English and 11 studies [40–50] published in Chinese. The sample size per study ranged from 28 to 1,591 and the total sample size was 8,080 (4,867 males, 3,213 females). Seven studies [18, 20, 21, 23, 29, 33, 44] were rated as high quality and 27 studies [17, 19, 22, 24–28, 30–32, 34–43, 45–50] were rated as moderate quality according to the NOS scale. The detailed characteristics and quality of the included studies are summarized in Table 1.

**Table 1.** Characteristics of included studies.

| Study | Year | Country | Language | Recruitment time frame | Sample | Age, years<sup>a</sup> | Sex |  | Cancer patients | NOS |
| --- | --- | --- | --- | --- | --- | --- | --- | --- | --- | --- |
|  |  |  |  |  |  |  | Male | Female |  |  |
| Chen NS [17] | 2020 | China | English | 2020.1.1-2020.1.20 | 99 | 55.5(13.1) | 67 | 32 | 1 | 6 |
| Chen T [18] | 2020 | China | English | 2020.1.13-2020.2.12 | 274 | 62(44-70) | 171 | 103 | 7 | 8 |
| Deng Y [19] | 2020 | China | English | 2020.1.1-2020.2.21 | 225 | 69(62-74)/<br>40(33-57)<sup>b</sup> | 124 | 101 | 8 | 6 |
| Feng Y [20] | 2020 | China | English | 2020.1.1-2020.2.15 | 476 | 53(40-64) | 271 | 205 | 12 | 8 |
| Grasselli G [21] | 2020 | Italy | English | 2020.2.20-2020.3.18 | 1591 | 63(56-70) | 1304 | 287 | 81 | 8 |
| Gautret P [22] | 2020 | France | English | 2020.3.3-2020.3.21 | 80 | 52.5(20-88) | 43 | 37 | 5 | 5 |
| Guan WJ [23] | 2020 | China | English | 2019.12.11-2020.1.31 | 1590 | 48.9(16.3) | 904 | 686 | 18 | 8 |
| Huang CL [24] | 2020 | China | English | 2019.12.16-2020.1.2 | 41 | 49(41-58) | 30 | 11 | 1 | 7 |
| Liu K [25] | 2020 | China | English | 2019.12.30-2020.1.24 | 137 | 57 (20-83) | 61 | 76 | 2 | 5 |
| Kim ES [26] | 2020 | Korea | English | 2020.2.1-2020.2.28 | 28 | 40(20-73) | 15 | 13 | 1 | 7 |
| Wang L [27] | 2020 | China | English | 2020.1.1-2020.2.6 | 339 | 69(65-76) | 168 | 171 | 15 | 6 |
| Mo PZ [28] | 2020 | China | English | 2020.1.1-2020.2.5 | 155 | 54(42-66) | 86 | 69 | 7 | 6 |
| Cai QX [29] | 2020 | China | English | 2020.1.11-2020.2.6 | 298 | 47.5 (33-61) | 145 | 153 | 4 | 8 |
| Shi HS [30] | 2020 | China | English | 2019.12.20-2020.1.23 | 81 | 49.5 (11.0) | 42 | 39 | 4 | 6 |
| Yang WJ [31] | 2020 | China | English | 2020.1.17-2020.2.10 | 149 | 45.1(13.4) | 81 | 68 | 2 | 7 |
| Wang DW [32] | 2020 | China | English | 2020.1.1-2020.1.28 | 138 | 56 (42-68) | 75 | 63 | 10 | 7 |
| Wu CM [33] | 2020 | China | English | 2020.12.25-2020.1.26 | 201 | 51 (43-60) | 128 | 73 | 1 | 8 |
| Wu J [34] | 2020 | China | English | 2020.1.22-2020.2.14 | 80 | 46.1(15.4) | 39 | 41 | 1 | 7 |
| Zhang XL [35] | 2020 | China | English | 2020.1.17-2020.2.8 | 645 | 34.9(14.2)/46.7(13.8)<sup>c</sup> | 328 | 317 | 6 | 6 |
| Yang XB [36] | 2020 | China | English | 2020.12.24-2020.1.26 | 52 | 59.7(13.3) | 35 | 17 | 2 | 7 |
| Zhang JJ [37] | 2020 | China | English | 2020.1.16-2020.2.3 | 140 | 57(25-87) | 71 | 69 | 6 | 7 |
| Zhou F [38] | 2020 | China | English | 2019.12.29-2020.1.31 | 191 | 56(46-67) | 119 | 72 | 2 | 6 |
| Zhu WB [39] | 2020 | China | English | 2020.1.24-2020.2.20 | 32 | 46(35-52) | 15 | 17 | 2 | 5 |
| Fang XW [40] | 2020 | China | Chinese | 2020.1.22-2020.2.18 | 79 | 45.1(16.6) | 45 | 34 | 1 | 6 |
| Gao T [41] | 2020 | China | Chinese | 2020.1.21-2020.2.18 | 40 | 41(16.4) | 19 | 21 | 2 | 5 |
| Huang LJ [42] | 2020 | China | Chinese | 2020.2.1-2020.2.24 | 71 | 41.3(16.7) | 38 | 33 | 2 | 5 |
| Li D [43] | 2020 | China | Chinese | 2020.1.20-2020.2.27 | 80 | 47.8(19.5) | 40 | 40 | 1 | 6 |
| Li YL [44] | 2020 | China | Chinese | 2020.2.5-2020.2.27 | 49 | 45(32-60) | 28 | 21 | 1 | 8 |
| Liu SJ [45] | 2020 | China | Chinese | 2020.1.23-2020.2.12 | 342 | 56(45-67) | 183 | 159 | 9 | 6 |
| Pan XQ [46] | 2020 | China | Chinese | 2020.2.5-2020.2.20 | 64 | 48.8(12.9) | 39 | 25 | 1 | 5 |
| Shi JH [47] | 2020 | China | Chinese | 2020.2.9-2020.2.29 | 54 | 62.5(50.5-68.5) | 31 | 23 | 10 | 6 |
| Wang R [48] | 2020 | China | Chinese | 2020.1.20-2020.2.14 | 96 | NR | 46 | 50 | 1 | 6 |
| Xiong J [49] | 2020 | China | Chinese | 2020.1.17-2020.2.20 | 89 | 53(16.9) | 41 | 48 | 11 | 5 |
| Zhang W [50] | 2020 | China | Chinese | 2020.1.21-2020.2.11 | 74 | 52.7(19.1) | 35 | 39 | 2 | 6 |

<sup>a</sup> Age data presented as median (IQR) or mean (SD); <sup>b</sup> data indicated death group/recovered group; <sup>c</sup> data indicated normal imaging findings group/abnormal imaging findings group. NR, not reported.

### 3.3. Prevalence of cancer in COVID-19 patients

All included studies [17–50] reported the number cancer patients among the COVID-19 patients. The meta-analysis showed that the pooled prevalence of cancer in patients with COVID-19 was 2.0% (95% CI: 2.0% to 3.0%), with significant heterogeneity among the studies (I^2^ = 68.8%, Figure 2). The Subgroup analysis indicated the prevalence was 5.0% (95% CI: 4.0% to 6.0%; 1 study [21], 1,591 patients) in Italy, 6.0% (95% CI: 3.0% to 14.0%; 1 study [22], 80 patients) in France, 4.0% (95% CI: 1.0% to 18.0%; 1 study [26], 28 patients) in Korea, and 2.0% (95% CI: 2.0% to 3.0%; 31 studies [17–20, 23–25, 27–50], 6,381 patients) in China (Figure 2).

Sensitivity analyses showed that the prevalence did not change after excluding studies with a sample size of less than 100 (Appendix Figure 1).

### 3.4. Association between cancer and the severity of COVID-19

Thirteen studies [20, 23, 24, 29, 32, 37, 40, 43–45, 47, 49, 50] involving 3,450 patients provided cancer data with comparison between COVID-19 patients with severe and non-severe illness. The meta-analysis revealed that cancer was associated with a 2.84-fold significantly enhanced risk of severe COVID-19 disease (OR = 2.84, 95%CI: 1.75 to 4.62, *P* < 0.001; I^2^ = 7.0%) (Figure 3). After excluding studies with a sample size of less than 100, we observed a stronger association (OR = 3.83, 95%CI: 2.21 to 6.63) between cancer and COVID-19 severity (Appendix Figure 2).

### 3.5. Association between cancer and the mortality of COVID-19

Six studies [18, 19, 23, 27, 36, 38], totaling 2,671 samples, reported cancer data between dead and surviving COVID-19 patients. Cancer was observed to be associated with a significantly enhanced risk of death (OR = 2.60, 95%CI: 1.28 to 5.26, *P* = 0.008; I^2^ = 6.2%), Figure 4. Sensitivity analysis by excluding studies with a sample size of less than 100 showed similar results (OR = 2.63, 95%CI: 1.14 to 6.06) (Appendix Figure 3).

### 3.6. Meta-regression analyses

Univariate meta-regression analyses revealed that publication languages and the number of centers of studies conducted were not the sources of heterogeneity or the factors affecting the cancer prevalence (Appendix Figure 4-5), the association between cancer and COVID-19 severity (Appendix Figure 6-7), and the association between cancer and the mortality of COVID-19 (Appendix Figure 8).

### 3.7. Publication bias

We found that there was a possibility of publication bias for cancer prevalence (*P* = 0.001) (Appendix Figure 9). Eggers tests indicated there was no statistically significant publication bias for the association between cancer and COVID-19 severity (*P* = 0.865) (Appendix Figure 10), and the association between cancer and COVID-19 mortality (*P* = 0.439) (Appendix Figure 11).

## 4. Discussion

### 4.1. Principal findings

This study included 34 studies from China, Italy, France, and Korea, identified from a comprehensive search of seven electronic databases. We systematically evaluated the prevalence of cancer among COVID-19 patients and the association between cancer and the severity and mortality of patients with COVID-19. Our meta-analyses indicated that the pooled prevalence of cancer in patients with COVID-19 was 2.0% and the prevalence in Italy, France, and Korea were higher than that in China. Cancer was associated with a 2.84-fold significantly enhanced risk of severe COVID-19 disease and a 2.60-fold significantly enhanced risk of death in patients with COVID-19. Sensitivity analyses showed that the results did not change substantially after excluding studies with a sample size of less than 100.

### 4.2. Comparison with other studies

A previous meta-analysis revealed that the pooled prevalence of malignancy among hospitalized COVID-19 patients was estimated to be 0.92% (95% CI: 0.56% to 1.34%) [51]. Another meta-analysis showed that the overall prevalence of cancer in patients with COVID-19 was 2.0% (95% CI: 2.0% to 3.0%), and the prevalence in studies with a sample size < 100 was slightly higher than that in studies with a sample size > 100 [52]. In the current study, we found the prevalence of cancer in COVID-19 patients was 2.0% (95% CI: 2.0% to 3.0%) and the prevalence did not change after excluding studies with a sample size of less than 100. Compared to these two studies, our study has several advantages that make it more conclusive. First, the present meta-analysis included 34 studies involving a total of 8,080 COVID-19 patients compared to no more than 3,561 patients in previous meta-analyses. Thus, our study had enlarged sample sizes and added statistical power of nearly 4500 cases. Second, studies included in previous meta-analyses were all from China, so the results may not apply to other countries and data selection bias may exist. However, our meta-analysis included studies from four countries, although only three studies from Italy, France, and Korea. Third, in addition to conducting subgroup analyses to evaluate the difference of prevalence between different countries, we also performed sensitivity analyses and meta-regression analyses and these analyses indicated that the results of our study were stable.

A previous meta-analysis found that there was no correlation between cancer and the severity of patients with COVID-19 [7]. However, our meta-analysis indicated that cancer was significantly associated with an increased risk of severe COVID-19 disease, which was inconsistent with the previous meta-analysis [7]. The previous reviewers conducted a meta-analysis based on only four studies with a sample of 1,356 patients [7]. We performed a meta-analysis of 3,450 cases from thirteen studies and sensitivity analysis by excluding studies with a sample size of less than 100 showed a stronger association. Therefore, the result of our study is more convincing.

### 4.3. Implications for research and practice

According to the current meta-analysis, the pooled prevalence of cancer in patients with COVID-19 was 2.0%. Combined with previously published results [9, 51, 52], we can conclude that patients with cancer have an increased risk of COVID-19. The development of cancer is usually related to a blunted immune status [53, 54], and anti-cancer treatments (such as chemotherapy and surgery) can also put cancer patients in an immunosuppressive state [8]. Therefore, immunodeficiency may be the main reason for cancer patients susceptible to COVID-19. Our subgroup analyses found that the prevalence of cancer among COVID-19 patients in Italy, France, and Korea were higher than that in China, although the result was limited by the sample size. These results suggest that cancer patients should be provided with special precautions and advised to use stronger personal protection.

Our meta-analysis found that cancer was associated with a 2.84-fold significantly increased risk of severe illness in patients with COVID-19, as well as with a 2.60-fold increased risk of death. Although our data are potentially limited by the sources of studies, these findings highlight the need for oncology professionals to be vigilant to the increased risk of serious illness and death associated with COVID-19 infection in patients with cancer or cancer survivors. A previous study showed that anti-tumor therapy within 14 days of COVID-19 diagnosis increased the risk of developing serious events and recommended that cancer patients with COVID-19 infection should avoid treatments causing immunosuppression [10]. However, there is currently no recommendation regarding the treatment strategies of immunotherapy, radiotherapy, chemotherapy, or delaying adjuvant therapy for cancer patients with COVID-19 [52]. The results of our meta-analysis also provide the latest references for the development of new guidelines. High-quality evidence-based guidelines clarify protection measures for cancer patients, and care and treatment strategies for cancer patients with COVID-19 are urgently needed.

### 4.4. Strengths and limitations

To the best of our knowledge, this is the first meta-analysis systematically evaluated the prevalence of cancer among COVID-19 patients, and the association between cancer and the severity and mortality of patients with COVID-19. Besides, we also conducted sensitivity analyses and meta-regression analyses to evaluate factors that may affect the results. However, our study also has some limitations. First, most of the studies included were from China, so the current findings may not fully reflect the global situation and should be interpreted with caution. Second, although this meta-analysis included 34 studies, there are few data available for subgroup analysis. Third, we performed sensitivity and meta-regression analyses to explore heterogeneity, but some factors were not evaluated due to limited data. Fourth, the patient overlap is still possible between a few studies, although we have ruled out many studies with overlap samples during the study selection and data extraction processes. Finally, we did not evaluate which types of cancer patients are more susceptible to COVID-19 or more associated with severe illness and mortality. As data from more countries become available, it is necessary to update this study and performed more comprehensive analyses to answer questions to guide clinical practice.

## 5. Conclusions

Our meta-analyses indicated that cancer patients have an increased risk of COVID-19, and cancer is associated with a 2.84-fold increased risk of severe illness and a 2.60-fold increased risk of death in patients with COVID-19. However, due to the limitations of this study, more high-quality studies from different countries are needed to provide robust evidence to support clinical practice.

## Data Availability

All data were provided in the manuscript.

## Abbreviations

COVID-19: Corona Virus Disease 2019; SARS-CoV-2: Severe acute respiratory syndrome coronavirus 2; COPD: chronic obstructive pulmonary disease; ARDS: acute respiratory distress syndrome; ICU: intensive care unit; PRISMA: Preferred Reporting Items for Systematic Reviews and Meta-Analyses; NOS: Newcastle-Ottawa quality assessment scale; OR: odds ratio; CI: confidence interval.

## Acknowledgments

The authors thank all investigators and supporters involved in this study.

## Authors contributions

YG, MLM, and JHT planned and designed the study. YG, ML, SZS, YMC, YS, and JC participated in the literature search and data collection. YG, ML, and SZS analyzed the data. YG and JHT drafted the manuscript. YG, ML, and JHT revised the manuscript. All authors read and approved the final manuscript.

## Funding

This work was supported by the Emergency Research Project of Key Laboratory of Evidence-based Medicine and Knowledge Translation of Gansu Province (Grant No. GSEBMKT-2020YJ01).

## Role of the Funding Source

The funders had no role in the design and conduct of the study; collection, management, analysis, and interpretation of the data; preparation, review, or approval of the manuscript; and decision to submit the manuscript for publication.

## Ethics approval and consent to participate

Not applicable.

## Consent for publication

Not applicable.

## Competing interests

The authors declare that they have no competing interests.

---

**Figure 2**. Pooled prevalence of cancer among patients with COVID-19.

**Figure 3**. Association between cancer and the severity of COVID-19 Figure 4. Association between cancer and the mortality of COVID-19

**Appendix Word 1**. Search strategy of PubMed.

**Appendix Figure 1**. Sensitivity analysis of cancer prevalence by excluding studies with a sample size of less than 100

**Appendix Figure 2**. Sensitivity analysis of the association between cancer and COVID-19 severity by excluding studies with a sample size of less than 100

**Appendix Figure 3**. Sensitivity analysis of the association between cancer and COVID-19 mortality by excluding studies with a sample size of less than 100

**Appendix Figure 4**. Univariate meta-regression analysis on the publication languages for the cancer prevalence

**Appendix Figure 5**. Univariate meta-regression analysis on the number of centers of studies for the cancer prevalence

**Appendix Figure 6**. Univariate meta-regression analysis on the publication languages for the the association between cancer and COVID-19 severity

**Appendix Figure 7**. Univariate meta-regression analysis on the number of centers of studies for the the association between cancer and COVID-19 severity

**Appendix Figure 8**. Univariate meta-regression analysis on the number of centers of studies for the the association between cancer and COVID-19 mortality

**Appendix Figure 9**. Funnel plot for cancer prevalence

**Appendix Figure 10**. Funnel plot for the association between cancer and COVID-19 severity

**Appendix Figure 11**. Funnel plot for the association between cancer and COVID-19 mortality

## Notes

### Competing Interest Statement

The authors have declared no competing interest.

## References

[1] Guan WJ, Ni ZY, Hu Y, Liang WH, Ou CQ, He JX, et al. Clinical Characteristics of Coronavirus Disease 2019 in China. The New England journal of medicine. 2020.

[2] Xu XW, Wu XX, Jiang XG, Xu KJ, Ying LJ, Ma CL, et al. Clinical findings in a group of patients infected with the 2019 novel coronavirus (SARS-Cov-2) outside of Wuhan, China: retrospective case series. BMJ (Clinical research ed). 2020;368:m606.

[3] WHO Director-Generals opening remarks at the media briefing on COVID-19: 11 March 2020. Published March 11, 2020. Accessed 22 April, 2020. https://www.who.int/dg/speeches/detail/who-director-general-s-opening-remarks-at-themedia-briefing-on-covid-19---11-march-2020.

[4] World Health Organization. Coronavirus disease (COVID-2019) situation reports. https://www.who.int/emergencies/diseases/novel-coronavirus-2019/situation-reports.

[5] Roncon L, Zuin M, Rigatelli G, Zuliani G. Diabetic patients with COVID-19 infection are at higher risk of ICU admission and poor short-term outcome. Journal of clinical virology: the official publication of the Pan American Society for Clinical Virology. 2020;127:104354.

[6] Hu Y, Sun J, Dai Z, Deng H, Li X, Huang Q, et al. Prevalence and severity of corona virus disease 2019 (COVID-19): A systematic review and meta-analysis. Journal of clinical virology: the official publication of the Pan American Society for Clinical Virology. 2020;127:104371.

[7] Wang B, Li R, Lu Z, Huang Y. Does comorbidity increase the risk of patients with COVID-19: evidence from meta-analysis. Aging. 2020;12:6049–57.

[8] Yang S, Zhang Y, Cai J, Wang Z. Clinical Characteristics of COVID-19 After Gynecologic Oncology Surgery in Three Women: A Retrospective Review of Medical Records. The oncologist. 2020.

[9] Liang WH, Guan WJ, Chen RC, Wang W, Li JF, Xu K, et al. Cancer patients in SARS-CoV-2 infection: a nationwide analysis in China. Lancet Oncology. 2020;21:335–7.

[10] Zhang L, Zhu F, Xie L, Wang C, Wang J, Chen R, et al. Clinical characteristics of COVID-19-infected cancer patients: A retrospective case study in three hospitals within Wuhan, China. Annals of oncology: official journal of the European Society for Medical Oncology. 2020.

[11] Moher D, Liberati A, Tetzlaff J, Altman DG. Preferred reporting items for systematic reviews and meta-analyses: the PRISMA statement. BMJ (Clinical research ed). 2009;339:b2535.

[12] Henry BM, de Oliveira MHS, Benoit S, Plebani M, Lippi G. Hematologic, biochemical and immune biomarker abnormalities associated with severe illness and mortality in coronavirus disease 2019 (COVID-19): a meta-analysis. Clin Chem Lab Med. 2020. doi: 10.1515/cclm-2020-0369.

[13] Lippi G, Wong J, Henry BM. Hypertension and its severity or mortality in Coronavirus Disease 2019 (COVID-19): a pooled analysis. Pol Arch Intern Med. 2020. doi: 10.20452/pamw.15272.

[14] Wells GA, Shea B, OConnell D, Peterson J, Welch V, Losos M, et al. The Newcastle-Ottawa Scale (NOS) for assessing the quality if nonrandomized studies in meta-analyses, 2012. Available from: http://www.ohrica/programs/clinicalepidemiology/oxfordasp.

[15] Zuin M, Rigatelli G, Zuliani G, Rigatelli A, Mazza A, Roncon L. Arterial hypertension and risk of death in patients with COVID-19 infection: systematic review and meta-analysis. J Infect. 2020. doi: 10.1016/j.jinf.2020.03.059.

[16] Higgins JP, Thompson SG, Deeks JJ, Altman DG. Measuring inconsistency in meta-analyses. BMJ (Clinical research ed). 2003;327:557–60.

[17] Chen N, Zhou M, Dong X, Qu J, Gong F, Han Y, et al. Epidemiological and clinical characteristics of 99 cases of 2019 novel coronavirus pneumonia in Wuhan, China: a descriptive study. Lancet (London, England). 2020;395:507–13.

[18] Chen T, Wu D, Chen H, Yan W, Yang D, Chen G, et al. Clinical characteristics of 113 deceased patients with coronavirus disease 2019: retrospective study. BMJ (Clinical research ed). 2020;368:m1091.

[19] Deng Y, Liu W, Liu K, Fang YY, Shang J, Zhou L, et al. Clinical characteristics of fatal and recovered cases of coronavirus disease 2019 (COVID-19) in Wuhan, China: a retrospective study. Chinese medical journal. 2020.

[20] Feng Y, Ling Y, Bai T, Xie Y, Huang J, Li J, et al. COVID-19 with Different Severity: A Multi-center Study of Clinical Features. American journal of respiratory and critical care medicine. 2020.

[21] Grasselli G, Zangrillo A, Zanella A, Antonelli M, Cabrini L, Castelli A, et al. Baseline Characteristics and Outcomes of 1591 Patients Infected With SARS-CoV-2 Admitted to ICUs of the Lombardy Region, Italy. Jama. 2020.

[22] Gautret P, Lagier JC, Parola P, Hoang VT, Meddeb L, Sevestre J, et al. Clinical and microbiological effect of a combination of hydroxychloroquine and azithromycin in 80 COVID-19 patients with at least a six-day follow up: A pilot observational study. Travel Med Infect Dis. 2020: 101663.

[23] Guan WJ, Liang WH, Zhao Y, Liang HR, Chen ZS, Li YM, et al. Comorbidity and its impact on 1590 patients with Covid-19 in China: A Nationwide Analysis. The European respiratory journal. 2020.

[24] Huang C, Wang Y, Li X, Ren L, Zhao J, Hu Y, et al. Clinical features of patients infected with 2019 novel coronavirus in Wuhan, China. Lancet (London, England). 2020;395:497–506.

[25] Liu K, Fang YY, Deng Y, Liu W, Wang MF, Ma JP, et al. Clinical characteristics of novel coronavirus cases in tertiary hospitals in Hubei Province. Chinese medical journal. 2020.

[26] Kim ES, Chin BS, Kang CK, Kim NJ, Kang YM, Choi JP, et al. Clinical Course and Outcomes of Patients with Severe Acute Respiratory Syndrome Coronavirus 2 Infection: a Preliminary Report of the First 28 Patients from the Korean Cohort Study on COVID-19. J Korean Med Sci. 2020;35:e142.

[27] Wang L, He W, Yu X, Hu D, Bao M, Liu H, et al. Coronavirus Disease 2019 in elderly patients: characteristics and prognostic factors based on 4-week follow-up. The Journal of infection. 2020.

[28] Mo P, Xing Y, Xiao Y, Deng L, Zhao Q, Wang H, et al. Clinical characteristics of refractory COVID-19 pneumonia in Wuhan, China. Clinical infectious diseases: an official publication of the Infectious Diseases Society of America. 2020.

[29] Cai Q, Huang D, Ou P, Yu H, Zhu Z, Xia Z, et al. COVID-19 in a Designated Infectious Diseases Hospital Outside Hubei Province, China. Allergy. 2020.

[30] Shi H, Han X, Jiang N, Cao Y, Alwalid O, Gu J, et al. Radiological findings from 81 patients with COVID-19 pneumonia in Wuhan, China: a descriptive study. The Lancet Infectious diseases. 2020;20:425–34.

[31] Yang W, Cao Q, Qin L, Wang X, Cheng Z, Pan A, et al. Clinical characteristics and imaging manifestations of the 2019 novel coronavirus disease (COVID-19):A multi-center study in Wenzhou city, Zhejiang, China. The Journal of infection. 2020;80.

[32] Wang D, Hu B, Hu C, Zhu F, Liu X, Zhang J, et al. Clinical Characteristics of 138 Hospitalized Patients With 2019 Novel Coronavirus-Infected Pneumonia in Wuhan, China. Jama. 2020.

[33] Wu C, Chen X, Cai Y, Xia J, Zhou X, Xu S, et al. Risk Factors Associated With Acute Respiratory Distress Syndrome and Death in Patients With Coronavirus Disease 2019 Pneumonia in Wuhan, China. JAMA internal medicine. 2020.

[34] Wu J, Liu J, Zhao X, Liu C, Wang W, Wang D, et al. Clinical Characteristics of Imported Cases of COVID-19 in Jiangsu Province: A Multicenter Descriptive Study. Clinical infectious diseases: an official publication of the Infectious Diseases Society of America. 2020.

[35] Zhang X, Cai H, Hu J, Lian J, Gu J, Zhang S, et al. Epidemiological, clinical characteristics of cases of SARS-CoV-2 infection with abnormal imaging findings. International journal of infectious diseases: IJID: official publication of the International Society for Infectious Diseases. 2020.

[36] Yang X, Yu Y, Xu J, Shu H, Xia J, Liu H, et al. Clinical course and outcomes of critically ill patients with SARS-CoV-2 pneumonia in Wuhan, China: a single-centered, retrospective, observational study. The Lancet Respiratory medicine. 2020.

[37] Zhang JJ, Dong X, Cao YY, Yuan YD, Yang YB, Yan YQ, et al. Clinical characteristics of 140 patients infected with SARS-CoV-2 in Wuhan, China. Allergy. 2020.

[38] Zhou F, Yu T, Du R, Fan G, Liu Y, Liu Z, et al. Clinical course and risk factors for mortality of adult inpatients with COVID-19 in Wuhan, China: a retrospective cohort study. Lancet (London, England). 2020;395:1054–62.

[39] Zhu W, Xie K, Lu H, Xu L, Zhou S, Fang S. Initial clinical features of suspected coronavirus disease 2019 in two emergency departments outside of Hubei, China. Journal of medical virology. 2020.

[40] Fang XW, Mei Q, Yang TJ, Zhang L, Yang Y, Wang YZ, et al. [Clinical characteristics and treatment strategies of 79 patients with COVID-19]. Chinese Pharmacological Bulletin. 2020: 453–9.

[41] Gao T, Xu YL, Zhang M, He XP, Chen JH. [Epidemiological and clinical characteristics of 40 patients with coronavirus disease 2019 outside Hubei]. Chinese Journal of Respiratory and Critical Care Medicine. 2020;19:148–53.

[42] Huang LJ, Chen FC, Jiang XQ, Li ZH, Wang W, Liu YW [Clinical characteristics and therapy of novel corona virus pneumonia: 71 cases retrospective analysis]. Central South Pharmacy. 2020.

[43] Li D, Long YZ, Huang P, Guo WL, Wu SH, Zhou Q, et al. [Clinical characteristics of 80 patients with COVID-19 in Zhuzhou City]. Chinese Journal of Infection Control. 2020. doi:10.12138/j.issn.1671-9638.20206514.

[44] Li YL, Shan NB, Sun W, Wang BG, Li DF. [Comparative Study for Clinical Features between COVID-19 Patients with Conventional Type and Heavy/Critical Type]. Practical Journal of Cardiac Cerebral Pneumal and Vascular Disease. 2020: 14–8.

[45] Liu SJ, Cheng F, Yang XY, He J, Li H, Zhang W, et al. [A study of laboratory confirmed cases between laboratory indexesand clinical classification of 342 cases with Corona Virus Disease 2019 in Ezhou]. Laboratory Medicine. 2020. DOI: 10.3969/j.issn.1673-8640.2019.00.000.

[46] Pan X, Hu Z. [Analysis on Chinese medical clinical characteristics of 64 patients with common type COVID-19]. Journal of Wenzhou Medical University. 2020. http://kns.cnki.net/kcms/detail/33.1386.R.20200324.1332.002.html.

[47] Shi JH, Wang YR, Li WB, Gang R, Liu X, Xu L, et al. [Digestive system manifestations and analysis of disease severity in 54 patients with corona virus disease 2019]. Chinese Journal of Digestion. 2020. doi:10.3760/cma.j.issn.0254-1432.2020.03.000.

[48] Wang R, Xie LL, Du P, Fan HQ, Song MY. [Clinical characteristics of 96 hospitalized patients with coronavirus disease 2019]. Chinese Journal of Respiratory and Critical Care Medicine. 2020;19:144–7.

[49] Xiong J, Jiang W, Zhou Q, Hu XQ, Liu CY. [Clinical characteristics, treatment, and prognosis in 89 cases of COVID J2019]. Medical Journal of Wuhan University. 2020. doi: 10.14188/j.1671J8852.2020.0103.

[50] Zhang W, Hou W, Li TZ, A.X L, Pan W, Jin RH, et al. [Clinical characteristics of 74 hospitalized patients with COVID-19]. Journal of Capital Medical University. 2020. doi: 10.3969/j.issn.1006-7795.2020.02.003.

[51] Emami A, Javanmardi F, Pirbonyeh N, Akbari A. Prevalence of Underlying Diseases in Hospitalized Patients with COVID-19: a Systematic Review and Meta-Analysis. Archives of academic emergency medicine. 2020;8:e35.

[52] Desai A, Sachdeva S, Parekh T, Desai R. COVID-19 and Cancer: Lessons From a Pooled Meta-Analysis. JCO global oncology. 2020;6:557–9.

[53] Schreiber RD, Old LJ, Smyth MJ. Cancer immunoediting: integrating immunity’s roles in cancer suppression and promotion. Science (New York, NY). 2011;331:1565–70.

[54] Xia Y, Jin R, Zhao J, Li W, Shen H. Risk of COVID-19 for patients with cancer. The Lancet Oncology. 2020;21:e180.

